# Forty-seven percent of pregnancies have stigmatizing language in their clinical notes in an electronic health record cohort

**DOI:** 10.64898/2026.07.27.26359049

**Authors:** Neha Simha, Hannah Takasuka, Li-Ching Chen, Umair Khan, Tomiko T. Oskotsky, Marina Sirota, John A. Capra, Irene Y. Chen

## Abstract

Stigmatizing language in medical documentation may reflect and perpetuate bias, but its prevalence in obstetrics has not been systematically quantified. We applied a keyword-guided BERT classifier to 640,345 obstetric notes from 26,178 pregnancies at an academic medical center. Stigmatizing language was detected in 47% of 26,178 pregnancies. Black pregnancies had significantly higher odds of stigmatizing language compared with Asian (aOR=1.5, p=3×10^-8^) or White (aOR=1.4, p=6×10^-6^). Indicated and spontaneous preterm births were also significantly associated with stigmatizing language compared to term (aORs=1.5, 1.2; p=7×10^-12^, 0.01). Pregnant individuals with only 12th-grade maternal education were more likely to experience stigma than those with college (aOR=1.5; p=4×10^-14^). These findings provide evidence of differences in clinical documentation across race, education levels, and clinical conditions. They also demonstrate how automated natural language processing can enable systematic monitoring of bias in healthcare language at scale.

## Introduction

Stigmatizing language reflects a provider’s implicit and negative, unfavorable, or biased view of a patient, often leading to disparities in care quality and health outcomes.^1–3^ Terms such as “noncompliant,” “difficult,” or other negatively framed characterizations can influence clinician perception, shape clinical decision-making, and potentially affect patient-clinician relationships.^4,5^ Furthermore, with the 21st Century Cures Act mandating patient access to clinical notes, documentation practices have come under greater scrutiny, as stigmatizing language visible to patients may erode trust and exacerbate medical mistrust among already marginalized populations.^1,6^ Lastly, as artificial intelligence is increasingly used to supplement or aid clinical documentation,^7^ preventing language models from unknowingly encoding stigma remains important to ensure equitable care.

Obstetric care represents a particularly critical setting for examining this phenomenon. Black women in the United States experience maternal mortality rates more than three times higher than White women (50 vs. 15 deaths per 100,000 live births as of 2023) and face similarly elevated risks of severe maternal morbidity.^8–10^ These stark disparities persist even after accounting for clinical comorbidities and socioeconomic factors, pointing to the role of structural racism, implicit bias, and communication patterns in perpetuating inequities.^10–12^

At large academic medical centers, stigmatizing language appears more frequently in clinical notes for marginalized populations, including Black patients.^2,4,13^ Qualitative studies and small-scale quantitative analyses have identified patterns wherein Black women receive more negative descriptors in their medical records compared to White women, and this differential documentation may be associated with worse patient experiences and outcomes.^14,15^ However, systematic measurement of stigmatizing language at scale has remained challenging due to the unstructured nature of clinical text, limited access to large datasets, and privacy constraints^1,3^. Recent advances in natural language processing (NLP) have begun to address these barriers. Transformer-based models such as ClinicalBERT, pretrained on a large corpora of medical text, can capture contextual meaning and distinguish stigmatizing from non-stigmatizing language even when identical keywords appear in neutral context^16–18^ These models have demonstrated superior performance on clinical NLP tasks compared to general-domain models.^16,19,20^ Despite these technical capabilities, few studies have applied such methods at scale within real-world health system data to systematically examine how stigmatizing language varies across patient demographics or in specific clinical contexts.^1,3^

Here, we applied a keyword-guided ClinicalBERT approach to identify stigmatizing language across 640,345 obstetric clinical notes from 26,178 deliveries in a pregnancy cohort at the University of California, San Francisco (UCSF). Our objectives were threefold: (1) to quantify the prevalence of stigmatizing language in obstetric documentation during pregnancy, (2) to examine whether stigmatizing language differed by maternal race/ethnicity, education level, age, and preterm birth outcome, and (3) to assess whether observed racial differences persisted after adjustment for clinical and sociodemographic factors. Given previous work, we hypothesized that Black and Hispanic pregnancies would have higher rates of stigmatizing language in their clinical notes compared to White and Asian pregnancies, independent of clinical characteristics. By leveraging the unique structure of electronic health records (EHRs), which integrate both structured clinical data and unstructured narrative text, this study provides a scalable framework for identifying and monitoring bias in clinical documentation, with direct implications for equity-focused interventions in maternal health care.

## Results

### Linking the UCSF Perinatal Database to clinical notes yields a dataset of 26,178 deliveries

To identify demographic and clinical factors associated with individuals who have stigmatizing language documented during obstetric care, we defined cohorts of preterm and term deliveries based on curated data from the UCSF Perinatal Database (PDB) and linked these to clinical notes from the UCSF EHR database. The cohort comprised 26,178 deliveries between 2012 and 2024, linked to 640,345 notes (Table 1).

**Table 1.** Maternal cohort characteristics of births at UCSF, 2012–2024.

| Characteristic | Value |
| --- | --- |
| Births, n | 26178 |
| Maternal age, years |  |
| Mean (SD) | 34 (5) |
| Median (IQR) | 34 (31-37) |
| Race/Ethnicity, n (%) |  |
| White | 12085 (46%) |
| Asian | 6536 (25%) |
| Hispanic | 3498 (10%) |
| Multiracial | 1429 (6%) |
| Black | 1384 (5%) |
| Unknown/ Declined | 827 (3%) |
| Other race | 290 (1%) |
| Pacific Islander | 86 (0.3%) |
| Native American | 43 (0.2%) |
| Obstetric History |  |
| Gravidity, median (IQR) | 2 (1-3) |
| Parity, median (IQR) | 1 (0-1) |
| Nulliparous, n (%) | 11741 (45%) |
| Gestational age, average weeks (SD) | 38 (3) |
| Preterm birth (<37 weeks), n (%) | 3,543 (14%) |
| Cesarean section, n (%) | 7471 (29%) |

### A ClinicalBERT classifier identified stigmatizing language in 47% of pregnancies

Stigmatizing language in medical documentation may reflect and perpetuate bias, but its prevalence in obstetric care has not been systematically quantified at scale. We applied a keyword-guided ClinicalBERT classifier to 640,345 obstetric clinical notes from 26,178 pregnancies at UCSF (Figure 1A). The classifier evaluated keywords spanning three categories: compliance (e.g., “nonadherence” and “refusal”), credibility/obstinacy (e.g., “adamant” and “claim”), and behavioral descriptors (e.g., “belligerent” and “drug seeking”), and classified each keyword instance as stigmatizing or non-stigmatizing based on surrounding context (Methods).

**Fig 1.**
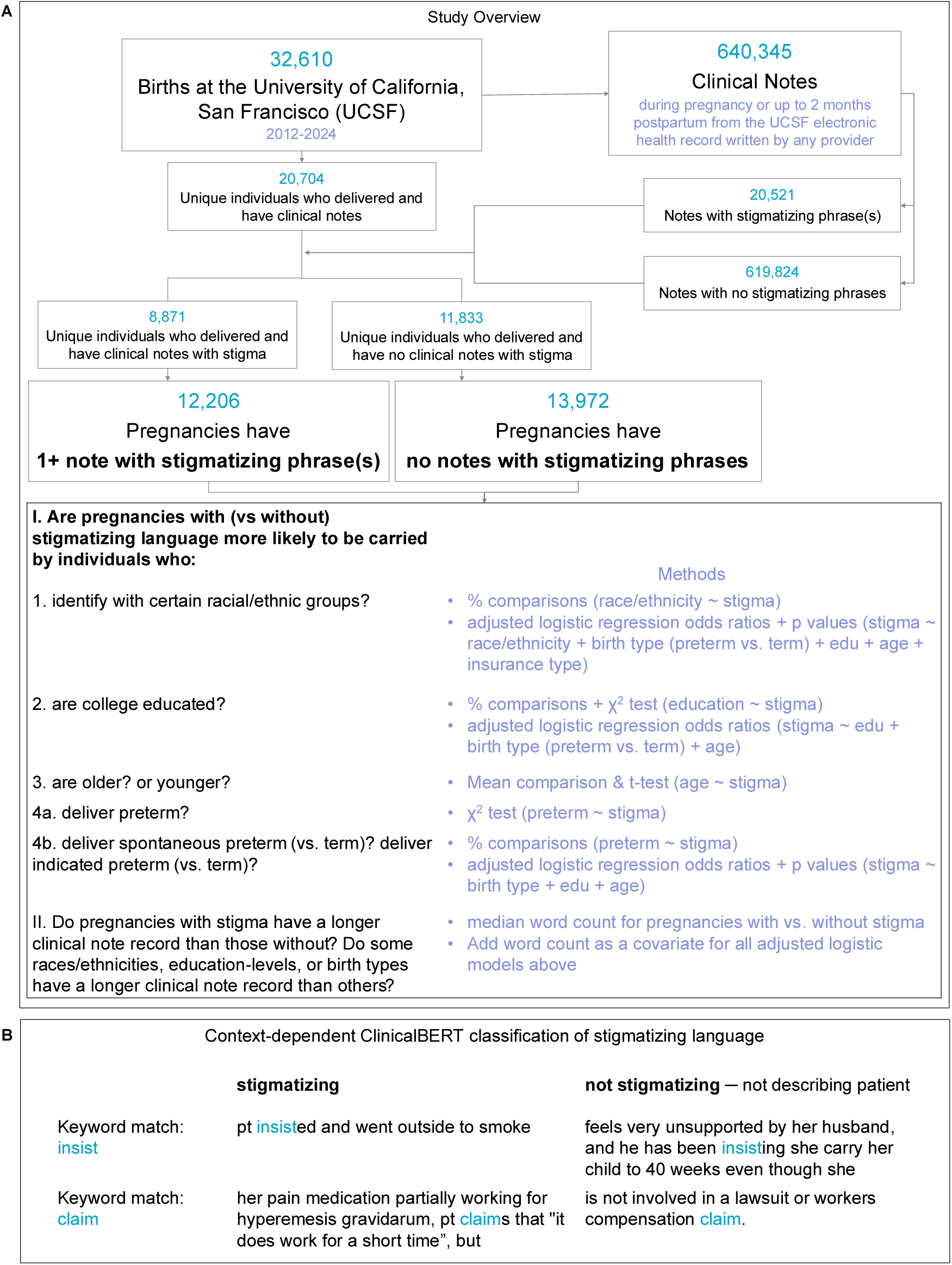
Overview of the study. (A) We quantify the use of stigmatizing language in the 640,345 clinical notes from 20,704 individuals and 26,178 pregnancies across demographics and clinical outcomes. (B) Clinical note excerpts illustrating keyword search and ClinicalBERT classification. Keyword search captures both stigmatizing and non-stigmatizing usage of terms like “insist” and “claim.” BERT classifies each instance based on the surrounding context, distinguishing non-stigmatizing clinical documentation (right examples) from potentially stigmatizing characterizations (left examples).

Across all notes, 20,521 (3%) contained at least one instance of stigmatizing language. At the pregnancy level, 12,206 of 26,178 pregnancies (47%) had at least one clinical note containing stigmatizing language. The contextual modeling of the provider’s description of the patient enabled our classifier to distinguish stigmatizing vs. non-stigmatizing uses of identical keywords (Figure 1B). This demonstrates the limitations of keyword search alone and the value of contextual modeling. The prevalence of stigmatizing language varied across the three semantic categories. Stigmatizing language was most common in compliance-related language (32% of pregnancies), followed by behavioral descriptors (22%) and credibility/obstinacy descriptors (1.1%).

### Manual review validates ClinicalBERT classification of stigmatizing language

To validate the accuracy of ClinicalBERT’s classification of stigmatizing language, we manually reviewed a sample of 100 notes that had a keyword and classified each note. We found high agreement (F1 = 0.89, Table 2). The 14 classification disagreements between BERT and manual review were adjudicated as follows: manual reviewer misclassified (n=4); BERT misidentified that the newborn or partner was the subject of the stigmatizing language and not the patient (n=3); subject is not explicit — BERT might have assumed the subject was not the patient while manual reviewer implied the subject to be the patient (n=3); the manual reviewer reads a positive or neutral description while BERT identified stigma (n=2); BERT sees stigma in an unchecked template checklist on whether the patient “refused blood products” while the manual reviewer does not (n=2). Thus, the algorithm performs well, and several of the cases of disagreement with manual review involve incomplete information or differences in interpretation, rather than clearly incorrect classifications.

**Table 2.** Validation of ClinicalBERT classification of stigmatizing language.

|  | Manual review -<br>not stigmatizing | Manual review -<br>stigmatizing |
| --- | --- | --- |
| BERT - not stigmatizing | 29 notes | 7 notes |
| BERT - stigmatizing | 7 notes | 57 notes |

### Black pregnancies had the highest prevalence of stigmatizing language

Racial disparities in healthcare quality and patient-provider interactions between Black and White patients are well-documented.^21–23^ There are fewer studies with Asian, Hispanic/Latine, and other individuals.^24^ Racial and ethnic disparities in clinical documentation have not been systematically examined in obstetric populations. We assessed the association between maternal race/ethnicity and exposure to stigmatizing language.

Stigmatizing language was unevenly distributed across maternal racial groups (Figure 2A, χ² = 135, df = 10, p = 4 × 10⁻²⁴, chi-squared test). Black pregnancies exhibited the highest prevalence of stigmatizing language (58%). White pregnancies, the largest racial group in the cohort, had a prevalence of 46%, and Asian pregnancies had a prevalence of 44%. The odds of stigmatizing language were significantly higher in the Black compared to White, even after adjusting for education level, insurance status, binary preterm vs. term clinical outcome, and maternal age (aOR = 1.5; p = 1 × 10⁻^10^). Black, multiracial, and White pregnancies also showed significantly elevated adjusted odds compared with Asian (Figure 2B). These disparities suggest that racial bias may influence clinical documentation practices in obstetric care.

**Figure 2.**
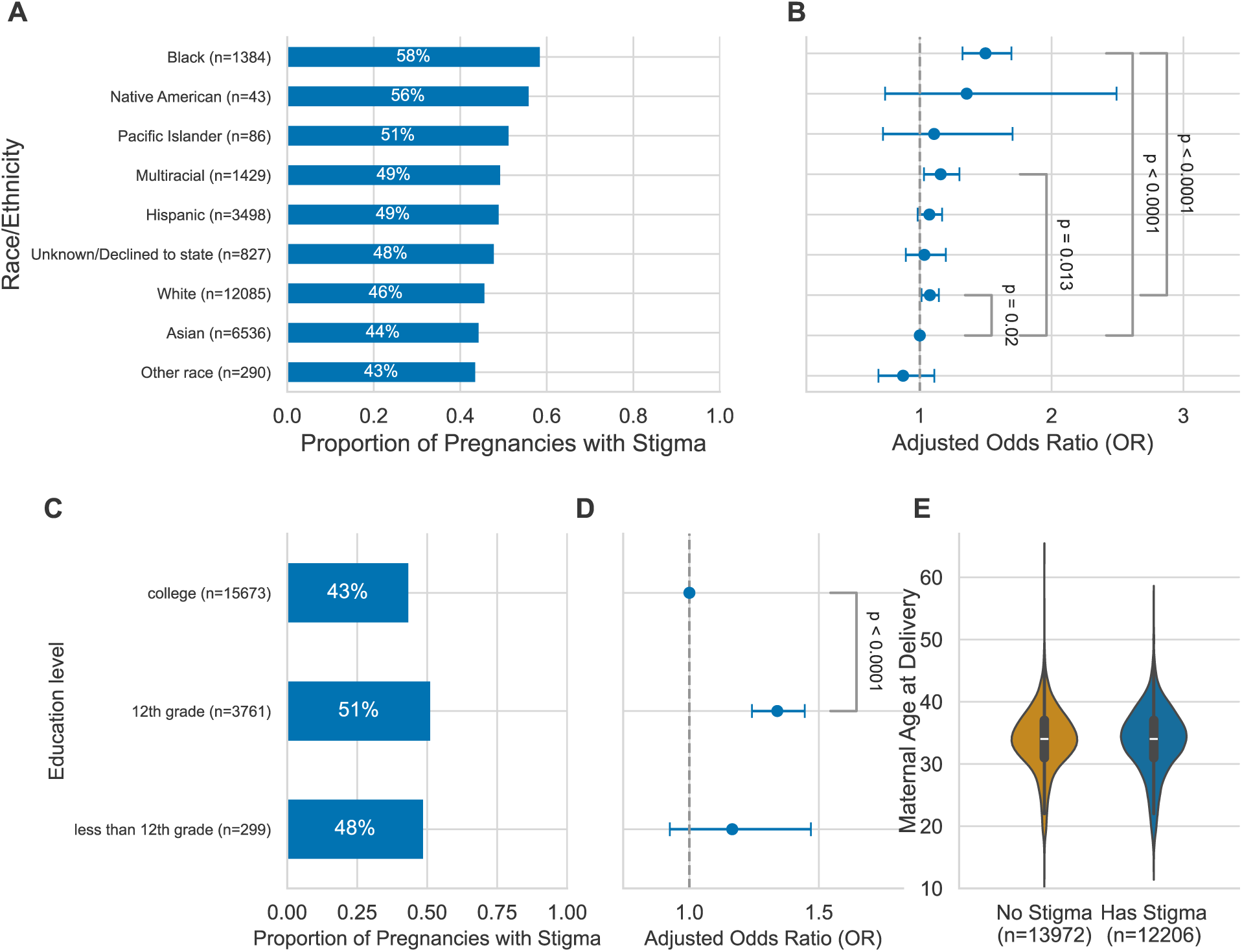
Stigmatizing language is more common in Black and lower education level pregnancies. (A) Proportion of pregnancies with at least one instance of stigmatizing language in their clinical notes, stratified by maternal race/ethnicity. (B) Adjusted odds ratios (aORs) and 95% confidence intervals for presence of stigmatizing language, estimated from multivariable logistic regression. Reference: Asian race. Covariates: preterm vs. term birth, maternal age, public vs. private insurance, maternal education level. (C) Proportion of pregnancies with at least one instance of stigmatizing language in their clinical notes, stratified by maternal education level. (D) The stigmatizing language odds differences between completing 12th-grade and completing college are significant after adjusting for binary preterm vs. term clinical outcome and maternal age. (E) Distribution of maternal age for pregnancies with vs without at least one clinical note containing stigmatizing language. Box plots show median and interquartile range.

### Less education associates with more stigmatizing language

Clinicians have been found to write stigmatizing notes that include stereotyping patient quotes with incorrect grammar or nonstandard, over-simplified medical terms.^25^ Therefore, we evaluated whether lower maternal education levels are associated with more stigmatizing language. Forty-three percent of pregnancies with maternal college education were exposed to stigmatizing language, compared to 48% of pregnancies with less than a high school diploma and 51% of pregnancies with 12th grade education (Figure 2C). We evaluated the associations between education level and exposure to stigmatizing language. Births by individuals with twelfth-grade educations were more likely to have stigma than births by individuals with college (aOR = 1.3, p = 5 × 10^-14^, Figure 2D) after adjusting for binary preterm vs. term clinical outcome and maternal age. Pregnancies with less than a 12th-grade maternal education had slightly higher odds of stigmatizing language than pregnancies with college (aOR = 1.2), though not significant (p = 0.2), likely due to small sample size. Lower education is generally associated with worse outcomes such as higher preterm birth,^26^ and stigmatizing language could play a role in potential causal mechanisms between education level and birth outcomes.

### Stigmatizing language is not clinically meaningfully associated with maternal age

Given that stigmatizing language might reflect ageist biases^27^ or differences in communication patterns across generations, we examined whether maternal age was associated with exposure to stigmatizing language. We compared maternal age distributions between pregnancies with and without stigmatizing language in their clinical notes. Maternal age distributions were nearly identical among pregnancies with and without stigmatizing language (Figure 2E). Mean maternal age differed by only 0.12 years between groups (33.79 vs. 33.67 years, t = 1.8, p = 0.06, two-sample t-test), with substantial overlap across the full age distribution. Thus, maternal age does not account for substantial variation in stigmatizing language exposure, suggesting that age-based biases do not substantially drive stigmatizing documentation in our cohort.

### Preterm births experience more stigmatizing language than term births

Preterm birth is a major adverse pregnancy outcome that may influence clinical documentation. Preterm births encompass heterogeneous clinical scenarios: spontaneous labor, membrane rupture, and medically indicated deliveries. These may relate differently to stigmatizing language. Using clinician curated annotations from the perinatal database, we categorized preterm births into three groups: (1) no preterm birth (term delivery ≥37 weeks, n = 22,635), (2) spontaneous preterm birth (spontaneous labor or preterm premature rupture of membranes, n = 1,288), and (3) indicated/iatrogenic preterm birth (medically indicated or iatrogenic delivery, n = 1,523). We excluded four pregnancies with preterm labor symptoms who delivered at term and 732 with unknown preterm status. We assessed associations between preterm category and stigmatizing language using chi-squared tests and pairwise logistic regression.

Overall, preterm birth was modestly and significantly associated with stigmatizing language (χ² = 101, df = 2, p = 9 × 10^⁻24^). Pregnancies resulting in indicated preterm births had a higher prevalence of stigmatizing language (57%) compared with term births (45%, aOR = 1.5, p = 1 × 10^-13^, logistic regression, Figure 3) after adjusting for maternal age, education level, and insurance status. Spontaneous preterm pregnancies also had a higher prevalence (49%) than term pregnancies (aOR = 1.2, p = 0.01, logistic regression). Thus, the relationship between preterm birth and stigmatizing language depends on the mechanism of preterm delivery. This suggests that clinical documentation may differ based on whether preterm birth reflects unanticipated events (spontaneous) versus provider decision-making (indicated).

**Figure 3.**
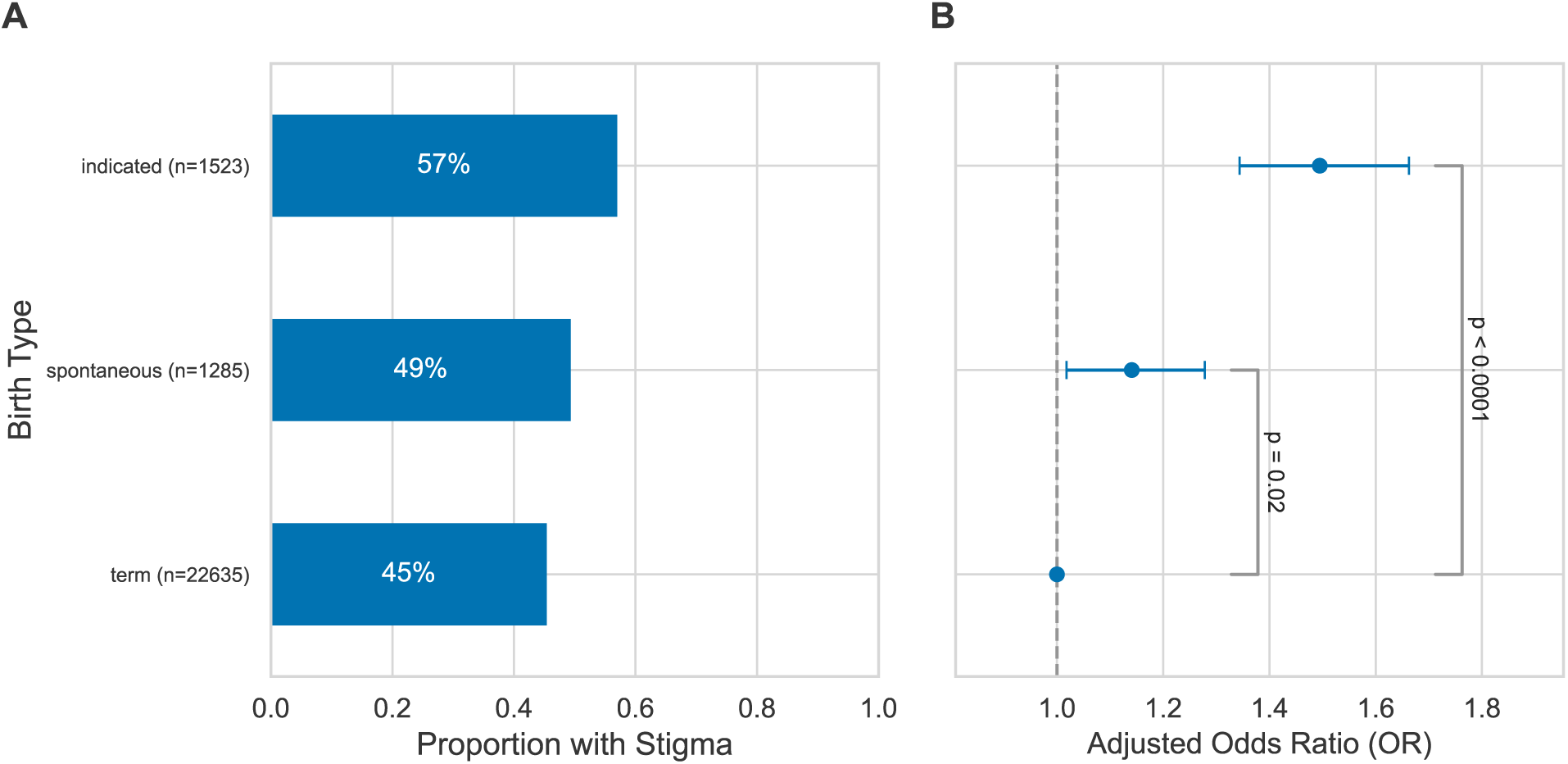
Pregnancies ending in indicated or spontaneous preterm birth are more likely to have stigmatizing language. (A) Proportion of pregnancies with at least one instance of stigmatizing language in their clinical notes, stratified by indicated preterm, spontaneous preterm, and term births. (B) Adjusted odds ratios and 95% confidence intervals for the presence of stigmatizing language, estimated from multivariable logistic regression. Covariates: maternal age, college education, public vs. private insurance. Reference: term birth.

### Black pregnancies are more likely to have stigmatizing language after adjusting for clinical note word count

Because we include pregnant individuals who receive full prenatal care at UCSF in addition to those who transferred or started care on delivery day, we hypothesize that pregnancies with a longer note record would be more likely to experience stigmatizing language. We evaluated the summed word count of all notes per pregnancy. Pregnancies with stigma had double the median word count (9,441 words) as pregnancies without (4,354 words, Figure 4A).

**Figure 4.**
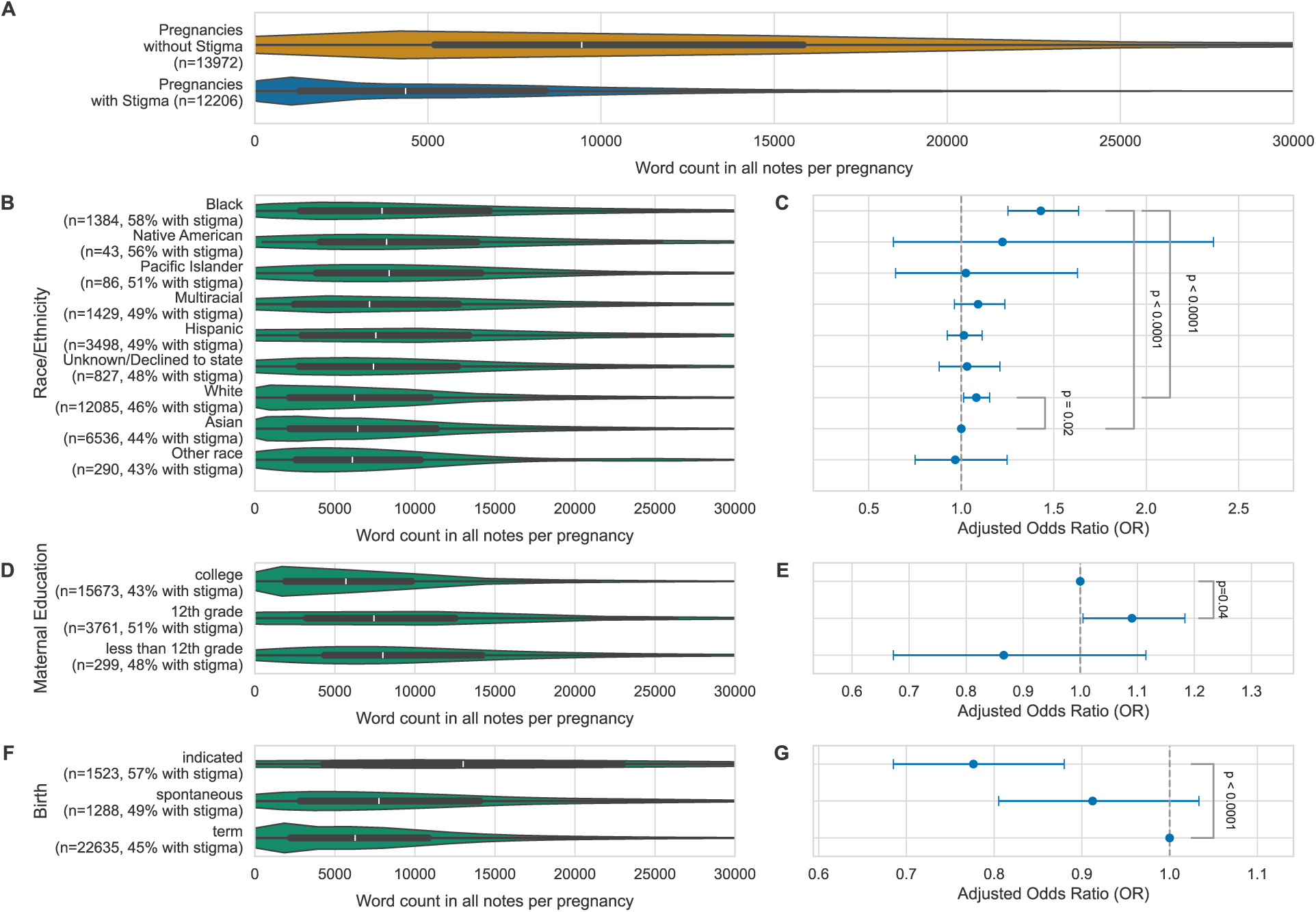
Pregnancies with stigma are more likely to have a longer clinical note record. (A) Distribution of word count in all clinical notes per pregnancy stratified by those with vs without at least one note containing stigmatizing language. (B) Distribution of word count per pregnancy, stratified by maternal race/ethnicity. (C) Adjusted odds ratios (aORs) and 95% confidence intervals for presence of stigmatizing language, estimated from multivariable logistic regression. Reference: Asian race. Covariates: word count, preterm vs. term birth, maternal age, public vs. private insurance, maternal education level. (D) Distribution of word count per pregnancy, stratified by maternal education level. (E) Pregnancies in twelfth-grade education level individuals have higher odds of stigmatizing language after adjusting for word count in addition to binary preterm vs. term clinical outcome and maternal age. (F) Distribution of word count per pregnancy, stratified by birth type. (G) Adjusted odds ratios and 95% confidence intervals for the presence of stigmatizing language, estimated from multivariable logistic regression. Covariates: word count, maternal age, college education, public vs. private insurance. Reference: term birth. (A, B, D, and F) Box plots show median and interquartile range; plot range is truncated at 30,000 words.

Given these differences, we evaluated whether note record length contributes to differences in stigma by race/ethnicity, maternal education, or birth type. To evaluate whether there are differences in note record length by demographic and clinical features, we plotted the distributions of word count stratified by race/ethnicity, maternal education, and birth type (Figure 4BDF). Noting many differences, we repeated all previous adjusted odds ratio analyses (Figures 2BD, 3B), adding word count as a covariate.

The median Black pregnancy had a 27% longer word count than White and 23% longer than Asian. After adjusting for the longer word counts, Black pregnancies have higher odds of stigma than White or Asian (aORs = 1.3,1.4; ps = 0.00002, 1 × 10^-7^, Figure 4C). This suggests that the higher prevalence of stigma in clinical notes of Black pregnancies is not due to longer records.

Pregnancies carried by individuals with a college education had a median 31% more words than those with a 12th-grade education. After adjusting for the longer note record, pregnancies carried by individuals with 12th-grade education only remain more likely to have stigma than those with college education (aOR = 1.1, p = 0.04, Figure 4E). However, the attenuation of the signal suggests that note length is a factor in the overall relationship between education level and stigma.

As expected based on the increased monitoring of at-risk pregnancies, pregnancies ending in indicated preterm birth had double the median number of words than term. Pregnancies ending in spontaneous preterm birth had a median 23% more words than those ending in term birth. These differences are large enough to change the direction of the odds ratios relating stigma and birth type. Indicated preterm births have significantly lower occurrence of stigmatizing language than term births when controlling for record word count (aOR = 0.8, p = 0.00007, Figure 4G). However, different clinical interactions may not have the same likelihood for stigmatizing language, e.g., stressful, high-risk communication during delivery vs. routine visits.

## Discussion

Despite growing recognition that language in medical documentation can reflect and perpetuate bias, tools to identify and quantify stigmatizing language at scale have been lacking. In this study, we demonstrate that automated natural language processing methods can systematically detect stigmatizing language in obstetric clinical notes. Applying a keyword-guided ClinicalBERT classifier across more than 640,000 clinical notes from 26,178 deliveries, we found that stigmatizing language is common. It affects 43-46% of pregnancies in White, Asian, or college-educated individuals, with 5-15% higher prevalence for pregnancies in individuals who are Black, deliver preterm, or have less education. Hispanic pregnancies were not more likely to contain stigmatizing notes than White or Asian. Overall, our findings provide quantitative evidence of racial, socioeconomic, and clinical inequities in clinical documentation and establish a scalable framework for measuring bias in healthcare language.

The persistence of Black racial disparities after adjustment for maternal age, maternal education, insurance status, preterm birth, and note length suggests that some differences in stigmatizing language cannot be explained by patient socioeconomic status or clinical risk alone. Instead, these disparities likely reflect systematic differences in how clinicians describe and characterize patients from different racial backgrounds. This finding aligns with extensive literature documenting racial bias in pain assessment^28,29^ and treatment recommendations^30^ and extends this work to demonstrate that bias is also embedded in written clinical narratives. The lack of associations with maternal age further supports the interpretation that stigmatizing language does not simply track with objective patient characteristics, but may reflect implicit racial bias, stereotyping, or differential framing of similar clinical presentations across racial groups.

Stratification of preterm birth revealed relationships between pregnancy outcomes and stigmatizing language. Previously, we found that medically indicated preterm births in our cohort have a higher prevalence of chronic conditions in comparison to term or spontaneous preterm births^31^. Combined with the results of this study, indicated preterm pregnancies represent the longest clinical note records with the most chronic conditions and the highest likelihood of experiencing stigmatizing language at least once. Spontaneous preterm pregnancies have a slightly higher prevalence of chronic conditions and slightly higher note records than term pregnancies. Spontaneous pregnancies are also slightly more likely to have at least one stigmatizing note than term. These findings demonstrate that stigmatizing language is more likely in pregnancies with complex clinical conditions.

This study has several limitations that future work should address. First, our analyses were conducted within a single tertiary care academic health system serving a diverse urban population, which may limit generalizability to other settings, particularly community hospitals or rural practices where patient demographics and documentation practices differ. Second, we quantified stigmatizing language as a binary outcome at the patient level, which may obscure important variation in frequency, severity, or the specific contexts in which it appears. Pregnancies with a single instance of stigmatizing language across dozens of notes were classified identically to those with pervasive stigmatizing documentation, potentially underestimating disparities. Third, while the ClinicalBERT model enables large-scale measurement and demonstrates strong performance in prior validation work and our validation, automated classification cannot fully capture clinician intent, tone, or the full spectrum of subtle language choices that may signal bias. Some instances of stigmatizing language may be missed, while others may be misclassified. Finally, our keyword-based approach limits detection to prespecified terms, potentially missing novel or emerging forms of stigmatizing language. Despite these limitations, our approach provides a scalable, reproducible framework for measuring bias in clinical documentation, offering a foundation for institutional monitoring and intervention evaluation.

AI scribes and other large language model (LLM) assistance in clinical note generation may alter the presence of stigmatizing language in clinical notes. Overall, a meta-analysis found that AI-generated clinical notes have similar quality to human-written notes.^32^ This study noted that AI-introduced inaccuracies are the largest concern. Inaccuracies, including hallucinations, accidental inclusions, and bias, may introduce stigmatizing language in some notes. As these AI scribe inaccuracies occur in over 10% of notes,^33^ it is important for clinicians to continue closely reviewing AI-generated clinical notes and correct inaccurate information. UCSF entered an AI scribe contract with Ambience Healthcare in December 2023;^34^ clinicians had the option to use this technology soon after, with patient consent. As our cohort represents deliveries 2012-2024, few clinical notes that we analyze in this study were written with AI LLM assistance.

Looking forward, this work opens several promising directions for research and practice. First, the methods developed here can be applied longitudinally to track temporal trends in stigmatizing language, assess the impact of bias training interventions, or evaluate policy changes aimed at promoting equitable care. Second, linking stigmatizing language to downstream patient outcomes such as treatment adherence, patient satisfaction, care continuity, or maternal morbidity would clarify whether biased documentation has tangible clinical consequences. Third, extending this framework to other clinical specialties or demographic dimensions (e.g., insurance status, body mass index, substance use history) could reveal broader patterns of bias in healthcare documentation. Fourth, examining clinician-level variation could identify specific documentation practices or training needs, enabling targeted interventions. We also encourage the development of real-time feedback tools that flag potentially stigmatizing language during documentation, providing clinicians with immediate opportunities to reflect on and revise their word choices. As healthcare systems increasingly adopt electronic health records and natural language processing, automated bias detection offers a tractable approach to monitoring equity and accountability in clinical communication. By making implicit biases visible and measurable, such tools can support institutional efforts to reduce disparities and center patient dignity in clinical care.

## Methods

### Data Source and Cohort Construction

We identified a pregnancy cohort using the UCSF perinatal database, which contains detailed demographic and clinical information on deliveries at UCSF Medical Center. The database includes 25,375 individuals who gave 32,610 births between June 1, 2012, and December 31, 2024. For each birth, we extracted maternal age at birth, self-reported race/ethnicity, gravidity, parity, gestational age, preterm birth category, and Cesarean section status.

This study and access to EHR data were approved by the Institutional Review Board of the University of California, San Francisco (#17–22929). As this was a retrospective study, patient consent was not applicable.

### Clinical Note Extraction and Preprocessing

We queried the UCSF clinical data warehouse^35^ to retrieve all clinical notes associated with individuals in the cohort. For each delivery, we included notes created within a perinatal window spanning 9 months before the delivery date through 2 months after delivery. This initial query yielded 2.67 million clinical notes across all individuals and deliveries.

We applied several preprocessing steps to focus on clinician-authored narrative documentation. First, we excluded all notes with missing or undefined note type designations. Second, we excluded rare note types (those with fewer than 100 total instances across the entire dataset) to ensure adequate representation for analysis. Third, we manually reviewed representative samples (minimum 20 notes per type) of the remaining note types to identify documentation formats likely to contain original clinician-written text. Notes consisting primarily of structured data, auto-populated templates, or patient-facing materials were excluded. Through this process, we retained 17 note types (Operative Report, Progress Notes, Lactation Note, H&P, RN Note, Consults, Discharge Summary, L&D Delivery Note, ED Provider Notes, Significant Event, Group Note, Consult, Care Coordination, ED Notes, ED Triage Notes, H&P [View-Only], and Student Note), resulting in a final analytic dataset of 640,345 clinical notes. These clinical notes mapped to 20,704 individuals who gave birth to 26,178 infants. We dropped deliveries without a clinical note.

### Keyword Identification and Span Extraction

We adapted a keyword-based detection approach from prior literature examining stigmatizing language in medical records^3^. From Harrigan et al., we obtained a comprehensive keyword list organized into three semantic categories: (1) credibility and obstinacy (e.g., “claims,” “adamant,” “denies”), (2) compliance (e.g., “noncompliant,” “nonadherent”), and (3) behavioral descriptors (e.g., “aggressive,” “difficult”). We searched for exact string matches of these keywords within the note text, treating keyword matching as case-insensitive.

For each keyword match, we extracted a contextual span that included the keyword and up to 10 tokens on either side. Tokens were defined using whitespace and punctuation boundaries. This span provided local context for downstream classification while limiting computational requirements. If a keyword occurred within 10 tokens of the note beginning or end, we extracted all available tokens within that boundary.

### ClinicalBERT Classification Model

We used a pretrained BERT model fine-tuned on clinical notes to classify each keyword span as stigmatizing or non-stigmatizing^3^. The base model (ClinicalBERT)^36^ was pretrained on clinical text from MIMIC-III. We applied a previously validated task-specific classification head to this model^3^.

For each extracted keyword span, we tokenized the text using the ClinicalBERT tokenizer and obtained contextualized embeddings from the final hidden layer of BERT. We applied anchor mean pooling to aggregate token embeddings corresponding to the keyword itself, creating a fixed-length representation. This pooled embedding was passed through a classification head consisting of a single linear layer with sigmoid activation, yielding a probability score for the stigmatizing class.

The classification model was trained and validated in prior work on manually annotated keyword spans from clinical notes^3^. The present study applies this trained model to our perinatal cohort without additional training.

### Stigmatizing Language Outcome Definition

We represented stigmatizing language as a multi-level binary outcome at three analytic levels: keyword, note, and pregnancy.

#### Identifying stigmatizing keywords

Harrigan et al. define that over 80 potentially stigmatizing keywords represent negative patient credibility/obstinacy, negative patient compliance, or negative patient behavioral descriptors. Each keyword instance was classified as stigmatizing if the BERT model assigned a probability ≥0.5 for the stigmatizing class. All other keyword instances (probability <0.5 or non-negative categories) were classified as non-stigmatizing.

#### Classifying stigmatizing notes

A clinical note was flagged as containing stigmatizing language if it contained at least one keyword instance classified as stigmatizing. Notes were assigned binary indicators for each of the three stigma categories (credibility/obstinacy, compliance, behavioral) as well as an overall “any stigma” indicator.

#### Identifying deliveries with individuals who experienced stigma

All deliveries by an individual were classified as having been exposed to stigmatizing language if any of the individual’s notes during any pregnancy episode contained stigmatizing language. Individual-level stigma indicators were constructed by taking the maximum probability of stigma across all notes associated with that individual for each stigma category.

### Preterm Birth Categorization

We categorized preterm birth into three clinically meaningful groups based on the mechanism of delivery as annotated by clinicians in the UCSF perinatal database.

- **No preterm**: Term delivery
- **Spontaneous**: Spontaneous preterm labor or Preterm premature rupture of membranes (PPROM)
- **Indicated/Iatrogenic**: Medically indicated preterm delivery, termination, or other iatrogenic preterm delivery

We excluded the four pregnancies with preterm labor with tocodynamometry (TOCO) monitoring but term delivery from the main analysis because these deliveries are clinically term births despite prodromal symptoms. We also excluded deliveries with unknown or missing preterm status.

### Statistical Analysis

#### Descriptive statistics

We computed frequencies and proportions for categorical variables (race/ethnicity, preterm type, maternal education level, stigmatizing language presence) and means with standard deviations for continuous variables (maternal age). All descriptive statistics were calculated at the individual level.

#### Bivariate associations

We assessed the association between stigmatizing language (binary outcome) and race using chi-squared tests. We similarly tested the association between stigmatizing language and birth type (term, spontaneous preterm, or indicated preterm) using chi-squared tests. For maternal age, we compared mean maternal ages between pregnancies with and without stigmatizing language using Welch’s t-test (unequal variances assumed).

#### Pairwise preterm comparisons

We performed post-hoc pairwise comparisons between preterm categories using Fisher’s exact test for each pair: (1) term vs. spontaneous, (2) term vs. indicated/iatrogenic, and (3) spontaneous vs. indicated/iatrogenic.

#### Multivariable logistic regression

We fit logistic regression models with presence of stigmatizing language as the binary outcome:

- **Binary preterm:** Predictors included race/ethnicity (categorical), maternal age (continuous), insurance type (private vs. public), summed word count of all notes (continuous), and preterm birth (binary: any preterm vs. none).
- **Stratified preterm:** Predictors included race/ethnicity (categorical), maternal age (continuous), insurance type (private vs. public), summed word count of all notes (continuous), and preterm category (categorical: term [reference], spontaneous, indicated/iatrogenic).

For each model, we computed adjusted odds ratios (aORs) with 95% confidence intervals and p-values for all predictor variables. We considered p < 0.05 statistically significant.

#### Software

All analyses were performed in Python 3.9.7. We used pandas (v1.4.2) for data manipulation, scipy (v1.9.0) for statistical tests, statsmodels (v0.13.2) for logistic regression, matplotlib (v3.5.2) and seaborn (v0.11.2) for visualization.

## Data and Code Availability

The UCSF perinatal database is maintained by the UCSF Maternal-Fetal Medicine Research team and contains protected health information. Access requires institutional approval and cannot be publicly shared. UCSF-affiliated individuals can request access to UCSF EHR clinical note data by contacting UCSF Information Commons.

The stigmatizing language ClinicalBERT model is publicly available at [https://www.github.com/kharrigian/ehr-stigma]. The keyword list is available at [https://www.github.com/kharrigian/ehr-stigma/blob/main/data/resources/keywords/keywords.json]. Analysis code is available upon request.

## Acknowledgements

We would like to acknowledge the T32 institutional training grant 5 T32DE0073-06, March of Dimes UCSF Prematurity Research Center, UCSF Lee Hysan Fund, Weill Cancer Hub West, Google Research Scholar award, and Apple Machine Learning grant for funding.

We would like to acknowledge Melissa Rosenstein for access to the Perinatal Database, the UCSF Information Commons team for EHR data access, and members of the Chen, Sirota and Capra Labs for useful discussion. We would like to thank Jean Costello for significant work in carrying out access requests to the Perinatal Database, linkage to EHR data, and associated data cleaning.

